# Factors influencing Blacks and Whites’ participation in Alzheimer’s disease biomarker research

**DOI:** 10.1101/2022.05.03.22274625

**Authors:** Johanne Eliacin, Elizabeth Hathaway, Sophia Wang, Caitlin O’Connor, Andrew J. Saykin, Kenzie A. Cameron

## Abstract

**INTRODUCTION:** Alzheimer’s disease (AD) is a public health priority. AD biomarkers may vary based on race, but recruitment of diverse participants has been challenging.

**METHODS:** Three groups of Black and White participants with and without prior research advocacy or participation were interviewed individually or in focus groups to better understand perspectives related to AD biomarker research participation. Thematic analytic approach was used to analyze the data.

**RESULTS:** Identified barriers to AD biomarker research participation included hesitancy due to fear, distrust of research and researchers, lack of relevant knowledge, and lack of research test results disclosure. Drivers for engagement in biomarker research procedures included knowledge about research, AD, and related clinical procedures, perceived benefits of participation, and outreach from trusted sources.

**DISCUSSION:** Participants’ comments related to the need for diversity in research and desire for results disclosure suggest opportunities to engage Black individuals.

## 1. Background

Over 6 million Americans are diagnosed with Alzheimer’s disease (AD), and this number is expected to increase to 12.7 million by 2050 (1). Given the high prevalence of AD and its associated financial, health, and societal impacts, research to reduce the disease burden of AD is a public health priority. The use of AD biomarkers [e.g., amyloid and tau cerebrospinal fluid (CSF) and imaging studies] to facilitate early detection and improved treatment of AD is currently a high priority research area. However, identification and interpretation of AD biomarkers are complicated by research findings indicating that they may vary based on race (2, 3). For example, several studies have suggested that Blacks may need different biomarker diagnostic thresholds because of lower CSF tau levels (4-6) compared to Whites. Other studies showed that the *APOE* gene seems to confer less AD risk among people of African ancestry relative to Whites (2, 3), suggesting a different AD disease process in this group.

Racial diversity among AD research participants is therefore essential to fully understand the role of biomarkers in AD and to ensure generalizability of AD studies. Such diversity is also important to reduce the disease burden for Black Americans, who are disproportionately impacted by AD with a prevalence of AD or other dementias roughly double that of White Americans (7, 8). However, recruitment of Black Americans for AD research studies has been a persistent challenge (9). Only 2.4% of participants in randomized controlled trials targeting cognitive function identified as Black (10). Yet they have been found to be as willing to participate in research as Whites (11).

Several factors have been identified to explain the low study participation rate among Black Americans. These include fear and mistrust of research based on a legacy of research misconduct, scientific exploitation, and racism (12-14). Other barriers include inequalities in healthcare systems and clinical trial designs that create financial, social, medical, and cultural barriers to research engagement (15-18).

Barriers and facilitators specific to AD biomarker research participation are, however, not well understood. Few studies have examined views of AD biomarker research participation among racially diverse groups (19)-(20). Biomarker research may involve additional barriers to participation given that it requires sharing of biological sample and undergoing relatively burdensome procedures such as lumbar puncture. Prior research suggested that Black participants are under-represented in drug trials and brain donation studies due to mistrust and concerns related to “mutilation or disfigurement of the body” (21). It is possible that these concerns also may apply to blood or CSF biomarker studies. With growing recognition of racial disparity in research participation and increased use of biomarkers in AD research, researchers are encouraged to take on a more active role in engaging racially diverse participants in research (22). In this study, we sought to examine participants’ perceived barriers to study participation and drivers of engagement in AD biomarker research, using semi-structured interviews, while seeking to identify differences between Black and White participants.

## 2. Methods

### 2.1 Setting and participants

This study was conducted in a midwestern city. Three groups of participants were invited to participate: Group 1: members of the Alzheimer’s Disease Research Center (ADRC)’s Community Advisory Board (CAB). The CAB includes community members, patients, and caregivers who support ADRC research; Group 2: community members and veterans from the local Veterans Health Affairs (VHA) Medical Center with no prior experience in AD research, and Group 3: ADRC research participants with normal cognition. Eligible participants identified as White or Black, were 55 years or older, and lived in the ADRC’s catchment area.

### 2.2 Recruitment and data collection

Participants were recruited either by direct outreach via e-mail, followed by a phone call, or with snow-balling techniques (23), i.e., asking enrolled participants to refer potentially eligible participants. Data were collected from January to March 2021. We conducted two 90-minute focus groups with CAB members; other study participants engaged in 45 to 60-minute semi-structured individual interviews. We used the framework of the Theory of Planned Behavior to develop our interview guide, which explored perceptions of AD research and assessed both drivers for engagement and barriers to participation in AD biomarker research studies (24). The interview guide was reviewed by two CAB members who did not participate in the focus groups. The first author (JE), who identifies as a Black female, facilitated the focus group discussions, while the study coordinator (CO), who is a White female, and two trained research assistants (one White and one Black, both female) in addition to JE conducted the individual interviews. Participants were compensated for their time and provided verbal consent prior to study participation. This study was approved by the university Institutional Review Board and the VHA Research and Development Review Committee.

#### 2.2.1 Data Analysis

Audio recordings of the focus groups and interviews were transcribed verbatim and reviewed for accuracy. A team of four analysts (JE, CO, and two research assistants) analyzed the data using a rapid data analysis approach, which involved summarizing the interview data into matrix summaries based on the interview guide and research questions (25, 26). We pilot tested the template with three transcripts and used team feedback to finalize it. Then, we generated a summary for each transcript. To ensure analytical rigor and trustworthiness, the team reviewed and discussed the content and summary of each transcript, resolving inconsistencies by consensus. We also established strategies such as peer debriefing meetings and audit trails to maintain consistency in our coding (27). The analytical team consolidated the interview summaries by domains and participant types (e.g., CAB, research participants) to identify commonly occurring themes, and to allow comparison across groups. We then identified broad themes and categories derived from the domains and conducted inductive thematic analysis (28) within each category, which were then reviewed with the larger research team.

## 3. Results

### 3.1 Participants

The study included 32 participants: 7 CAB members, 5 veterans, 10 ADRC participants, and 11 community members. As shown in Table 1, among the community and ADRC participants, fifteen were female (57.7%), all were non-Hispanic, and fifteen (57.7%) self-identified as Black or African American. Forty-two percent (41.7%) were 60-69 years old. Most (68.2%) had completed at least a four-year college degree. We did not collect demographic data from ADRC CAB members.

Overall, participants identified four key barriers to AD biomarker research participation: 1) hesitancy due to fear, 2) hesitancy due to distrust of research and researchers, 3) lack of knowledge about research, AD, and related clinical procedures, and 4) lack of research test results disclosure. In turn, drivers for engagement in biomarker research procedures included 1) knowledge of research, AD, and related clinical procedures, 2) perceived benefits to individual and community, and 3) outreach from trusted sources. Below we expand on each of these themes, noting differences identified across participant groups and based on race.

### 3.2 Barriers to AD Biomarker Research Participation

#### 3.2.1 Hesitancy due to fear

Hesitancy due to fear was the most frequently reported barrier to AD biomarker research across all participant groups. Identified fears included fear of the unknown about research and fear of a potential AD diagnosis, as illustrated by the following quote from a White female participant (205): “*I think it would be scary knowing for sure if I have the markers …You know, if I can’t find my car keys, then I’ll be sure that it’s beginning*.*”*

#### 3.2.2 Hesitancy due to distrust of research and researchers

Participants reported distrust in AD biomarker research, with variation in contributors to distrust noted based on race. Specifically, Black participants emphasized *lack of research transparency* as a key barrier to AD research participation and maintained that detailed information should be shared with participants during the early phase of the study to demystify the research process and develop trust. Participants also reported distrust in research due to *data safety and confidentiality concerns*. They explained that data collected from AD biomarker studies, such as genetic information, are sensitive, reflecting that any breach of confidentiality would have significant social, health, and legal implications not only for them personally, but also for their families. For example, one Black male veteran shared: “*I wouldn’t want to have the image done of my brain, and it being released to the insurance companies without my knowledge*” (403).

Both White and Black participants discussed how *racial bias and healthcare disparities* undermine trust in research. However, this theme was particularly salient in Black participants’ interviews. Many drew upon historical cases as well as their lived experiences and news reports of racial bias in healthcare to inform their opinions about AD biomarker research participation. As in the next excerpt, they questioned whether research can be truly benevolent towards communities of color and underscored the “*hurt” -* the emotional aftermath of racial bias and resulting stigma that undermine community support for research.

> *There is a hurt and a stigma in the Black community. They were guinea pigs for research. And that has come through generations of Black people, those stories are told through our community*.– Black female participant (311).

#### 3.2.3 Lack of knowledge about research, Alzheimer’s disease, and related clinical procedures

Both Black and White participants reported that limited knowledge about research and its value is a key barrier to research participation. They noted that this lack of knowledge may lead to disinterest and apathy in the research enterprise. To illustrate, a Black female ADRC participant stated: “*I would say a lack of education in that area (is a barrier) and that they [Black participants] probably don’t feel like doing it is going to make a difference”(209)*.

Participants also discussed how limited knowledge about the AD biomarker clinical procedures, such as brain imaging and PET scan, may deter potential participants from engaging in research. Additionally, several stated that older adults often have medical conditions and are hesitant to undergo unnecessary procedures that may exacerbate those conditions or put them at additional health risks.

#### 3.2.4 Lack of research test results disclosure

For most ADRC participants, previous ADRC research experiences when they did not receive test results was viewed as a barrier to future research participation and study retention:

> *I think any part of the study that do you, you ought to receive feedback because that’s going to encourage people to continue … when we go through those sessions, but we don’t hear results back, that causes me to not want to do the extra because the reason I’m participating in this study is to help give you information. And part of that information I want to know myself personally. So, that’s an important aspect. -*Black female ADRC participant (206)

CAB members and study participants with no prior research experience expressed similar views. Participants conveyed their perception that failure to receive test results, including normal results, can serve as a barrier to AD biomarker research participation: *“I think people need to be made aware of [test results]*.. *[if] they’re not going to know … that will make people hesitant*.” - Black male CAB member (107)

### 3.3 Common Facilitators/ Motivators to AD Biomarker Research Participation

Participants identified several drivers for engagement in AD biomarker research studies. Although we asked participants during the interviews about their perspectives about each biomarker study procedure, such as blood drawn for genetic testing, brain MRI, PET scans, and lumbar puncture, their responses were consistent regardless of type of study procedure.

#### 3.3.1 Knowledge of research, Alzheimer’s disease, and clinical procedures

Participants explained that prior experience with specific clinical procedures and/or research knowledge is a strong driver for future research participation. Many participants who expressed interest in participating in AD biomarker research reported prior positive research experiences, familiarity with the clinical procedures, and/or a healthcare background: *“I had a PET scan recently, of my abdomen. So, it would be the same experience. I would be willing to do that because I have tried it …. I can handle that*.*” White Male Community participant (306)*

#### 3.3.2 Perceived benefits to individuals and community

Participants reported that perceived benefits from AD biomarker studies are drivers for their engagement in research. Specifically, they identified benefits at both individual and community levels that contribute to their willingness to engage in research.

> *Individual benefits –* Most Black and White participants reported having a family member or friend with a history of AD; this personal connection to AD appeared to be a significant motivator for research participation. They expressed interest in AD biomarker research because of the potential of learning about their family history and health status, and gaining insights into how they can reduce their risks of developing AD.

> *My mother had early onset Alzheimer’s. … she was 57…[research participation] it’s just important to me because I’m certainly not out of the woods, and none of my kids, and for anybody who, and their caregivers who might be affected, I just want to help in any way I can. …if my mother had not had this disease, I doubt that I would have even thought about volunteering. White Female ADRC (206)*

*Community benefits* – Black and White participants, including those with no prior history of AD research participation, also reported without probing that they are interested in research involvement because of their desire to increase research diversity. However, this theme was most salient among Black participants. Many Black participants cited well-documented disparities in research and healthcare. As exemplified in the excerpt below, they stated they want to do their part to ensure that AD research studies are generalizable and equitable.

> *Because people of color are consumers of medicine, we’re going to need that [knowledge]. And if you’ve only tested something [on] a White male between the ages of 18 and 25, what does that mean for my mother or my grandmother, who didn’t fit into that category? So, I’ve always felt very strongly about representation… that’s the primary reason [for research participation]*. -Black female ADRC participant (201)

Moving beyond discussions about disparities in research and individual research incentives, Black participants specifically emphasized the need of research projects to address how the studies benefit Black communities. They advocated for early engagement of community members to create buy-in for the studies and for researchers to share the “wealth” – as in knowledge and resources with Black communities to facilitate trust and meaningful engagement.

> *A lot of times we’re not at the table*… *But if there is an understanding from the beginning all the way through, then I think you have more buy in, you have people that are more committed to it. And then they can talk from a place of how important and valuable it is for a person to be involved in that research*. – Black CAB Member

#### 3.3.3 Outreach from trusted sources

Personal outreach from trusted sources was identified as a major driver for engagement across all study groups. Participants underscored that learning about AD biomarker research studies from individuals with lived experiences as research participants would be most helpful. Black participants further emphasized the need for research team members to reflect the sociocultural and racial backgrounds of their participants to facilitate trust. Some specifically discussed the need for the entire team, including investigators, to reflect the diversity of the community and maintained that researchers must personally engage with potential participants to facilitate trust. A Black female CAB member explained:

> *There’s no way that I think anyone would have surgery if you just talked to a nurse, to a secretary…and didn’t meet that doctor. I don’t know why in a research situation the research doctor doesn’t think a person wants to talk to him or her. I want to talk to the person that’s doing it on me. … I need to eyeball you. I need to see if we can connect. If I can’t connect with you in conversation, I can’t allow you to experiment on my body*.*”*

## 4. Discussion

Our study identified hesitancy due to fear, distrust, and lack of knowledge about research as barriers to AD biomarker research participation. These findings are consistent with previous reports of barriers to AD biomarker research among racially diverse groups (19, 20). For example, Williams et al. (20) wrote more than a decade ago about how mistrust in and limited knowledge of research were fundamental reasons for nonparticipation in AD biomarker research among Black Americans. Many of these barriers such as lack of knowledge of AD are modifiable factors. Yet, they continue to persist. A recent Alzheimer’s Association study reported that 62% of Black Americans believe that medical research is biased against people of color (1). Together, these findings point to the continuing need for more effective recruitment and outreach strategies, and better implementation of evidence-based engagement methods. Intentional and sustained efforts to engage research participants also may help to address barriers related to confidentiality, transparency, and trust in research.

Our findings also highlight participants’ discussion about AD biomarker test results disclosure as a tool for research recruitment and engagement. Most participants identified non-disclosure of both normal and abnormal results as a significant barrier to research participation and stated their belief that sharing of the results may drive engagement in AD biomarker studies. These findings are noteworthy for several reasons. Previous studies on disclosure of genetic risks in research studies have focused almost exclusively on White participants (29, 30). Our findings add the perspectives of Black participants to this important discussion. Our findings also show that disclosure of research test results is a critical issue that may impact active participants’ retention and participation in future studies.

Moreover, research centers vary widely in their practice of AD research test results disclosure to study participants, with biomarker data less commonly disclosed than cognitive test results (31). Researchers have raised concerns regarding the potential psychological impact of disclosure, particularly for those receiving news of increased AD risk in the absence of a broadly available disease-modifying treatment. However, some studies also suggest that the risk of psychological harm is relatively low, especially with the provision of genetic counseling (30, 31, 33, 34), and efforts are underway to optimize disclosure protocols for dementia risks to research participants (32). While our findings indicate some support for sharing research test results with study participants, they also point to emerging views about the type of data participants want disclosed, such as both normal and abnormal results as well as non-genetic test results. Disclosure of test results with research participants undoubtedly raise ethical, financial, and procedural concerns for research institutions. However, as our findings indicate, it may also create opportunities for meaningful engagement with underserved communities and opportunities for community-wide health promotion.

Further, our findings expand on discussions of health disparities as barriers to AD research participation. They showed that ongoing reports of healthcare disparities and personal, lived experiences of racial bias in medicine influence participants’ perspectives of research participation. An implication of this finding is that research engagement efforts must address not only the historical legacies of racial discrimination in research but also individuals’ present and personal experiences of racial bias in health services.

Outreach from a trusted source was also reported as a motivator for engagement. Specifically, participants expressed wishes to engage with principal investigators prior to consenting to a study, suggesting that such engagement would provide greater research transparency and could instill trust among participants. Moreover, they identified research participants themselves as a potential powerful source for engagement given their lived experiences with AD biomarker research. Adapting a peer support (33, 34) or peer navigator (35-39) model that involves hiring individuals with lived research experiences to facilitate research outreach, education, and engagement in AD biomarker is a strategy to address these suggestions.

Several limitations should be considered in interpreting the results of this study. This study included mostly participants with high educational attainment, from one midwestern city. As such, responses may not represent the full range of perspectives of patients eligible for biomarker studies. The use of three distinct participant groups sought to capture a varied sample, but the perspectives obtained on research participation carry some inherent bias because these voices represent a group agreeable to at least a low-risk form of research engagement. Also, participants’ perspectives may not reflect their future behaviors. Demographic data was not collected from focus group participants to limit concerns about self-disclosure that might have interfered with engagement during the group interview. Despite these limitations, the study offers several contributions and incorporates various strengths such as inclusion of diverse participant groups with and without previous research experience. Building on our findings, future studies should further assess sociocultural factors that may impact diverse groups’ views of AD biomarker research participation. Development and testing of targeted strategies to meet diverse groups’ needs, and to address their specific barriers to AD biomarker research participation are warranted.

In summary, as with medical decision-making, personal decisions about research participation often hinge on a perceived risk-benefit analysis. Our findings illustrate how one’s individual experience may influence identified risks and benefits of AD biomarker participation. Culturally attuned engagement strategies may amplify those perceived benefits and diminish the perceived risks by building knowledge and bridging the barriers that separate the research community from potential participants.

## Data Availability

All data produced in the present work are contained in the manuscript.

## 5. Acknowledgements

The authors greatly acknowledge the support and contributions of the Indiana Alzheimer’s Disease Research Center Community Advisory Board in this project. They also thank the study participants who made this research possible.

## 6. Conflicts

None of the investigators have a conflict of interest. Dr. Wang receives book royalties from APPI and DSMB consultant fees (total less than $2000/year). Dr. Saykin receives support from Avid Radiopharmaceuticals, a subsidiary of Eli Lilly (in kind contribution of PET tracer precursor); Bayer Oncology (Scientific Advisory Board); Eisai (Scientific Advisory Board); Siemens Medical Solutions USA, Inc. (Dementia Advisory Board); Springer-Nature Publishing (Editorial Office Support as Editor-in-Chief, Brain Imaging and Behavior). Dr. Cameron receives consultant fees from EPI-Q, Inc.

## 7. Funding sources

This research was supported, in part, by grants from the National Institutes of Health, National Institute on Aging, [P30 AG10133, P30 AG072976, R01 AG019771] to AJS. Partial support for this work was also provided by a Veterans Health Affairs, Health Services Research and Development Career Development Award [16-153] and an Academy of Communication in Health Putnam Scholar Fellowship to JE. Partial support for this work was also provided by the Alzheimer’s Association LDRFP-21-818464 to SW and JE.

## Notes

### Competing Interest Statement

The authors have declared no competing interest.

### Author Declarations

The IRB of the Indiana University School of Medicine gave ethical approval for this work.

## References

1. 2021 Alzheimer’s disease facts and figures. Alzheimers Dement. 2021;17(3):327–406.

2. Farrer LA, Cupples LA, Haines JL, Hyman B, Kukull WA, Mayeux R, et al. Effects of age, sex, and ethnicity on the association between apolipoprotein E genotype and Alzheimer disease. A meta-analysis. APOE and Alzheimer Disease Meta Analysis Consortium. JAMA. 1997;278(16):1349–56.

3. Tang MX, Stern Y, Marder K, Bell K, Gurland B, Lantigua R, et al. The APOE-epsilon4 allele and the risk of Alzheimer disease among African Americans, whites, and Hispanics. JAMA. 1998;279(10):751–5.

4. Morris JC, Schindler SE, McCue LM, Moulder KL, Benzinger TLS, Cruchaga C, et al. Assessment of Racial Disparities in Biomarkers for Alzheimer Disease. JAMA Neurol. 2019;76(3):264–73.

5. Howell JC, Watts KD, Parker MW, Wu J, Kollhoff A, Wingo TS, et al. Race modifies the relationship between cognition and Alzheimer’s disease cerebrospinal fluid biomarkers. Alzheimers Res Ther. 2017;9(1):88.

6. Chaudhry A, Rizig M. Comparing fluid biomarkers of Alzheimer’s disease between African American or Black African and white groups: A systematic review and meta-analysis. J Neurol Sci. 2021;421:117270.

7. Rajan KB, Weuve J, Barnes LL, McAninch EA, Wilson RS, Evans DA. Population estimate of people with clinical Alzheimer’s disease and mild cognitive impairment in the United States (2020-2060). Alzheimers Dement. 2021;17(12):1966–75.

8. Gurland BJ, Wilder DE, Lantigua R, Stern Y, Chen J, Killeffer EH, et al. Rates of dementia in three ethnoracial groups. Int J Geriatr Psychiatry. 1999;14(6):481–93.

9. Shavers-Hornaday VL, Lynch CF, Burmeister LF, Torner JC. Why are African Americans under-represented in medical research studies? Impediments to participation. Ethn Health. 1997;2(1-2):31–45.

10. Vyas MV, Raval PK, Watt JA, Tang-Wai DF. Representation of ethnic groups in dementia trials: systematic review and meta-analysis. J Neurol Sci. 2018;394:107–11.

11. Wendler D, Kington R, Madans J, Van Wye G, Christ-Schmidt H, Pratt LA, et al. Are racial and ethnic minorities less willing to participate in health research? PLoS Med. 2006;3(2):e19.

12. Gamble VN. Under the shadow of Tuskegee: African Americans and health care. Am J Public Health. 1997;87(11):1773–8.

13. Hughes TB, Varma VR, Pettigrew C, Albert MS. African Americans and Clinical Research: Evidence Concerning Barriers and Facilitators to Participation and Recruitment Recommendations. Gerontologist. 2017;57(2):348–58.

14. Scharff DP, Mathews KJ, Jackson P, Hoffsuemmer J, Martin E, Edwards D. More than Tuskegee: understanding mistrust about research participation. J Health Care Poor Underserved. 2010;21(3):879–97.

15. Luebbert R, Perez A. Barriers to Clinical Research Participation Among African Americans. J Transcult Nurs. 2016;27(5):456–63.

16. Lincoln KD, Chow T, Gaines BF, Fitzgerald T. Fundamental causes of barriers to participation in Alzheimer’s clinical research among African Americans. Ethn Health. 2021;26(4):585–99.

17. Awidi M, Al Hadidi S. Participation of Black Americans in Cancer Clinical Trials: Current Challenges and Proposed Solutions. JCO Oncol Pract. 2021;17(5):265–71.

18. Otado J, Kwagyan J, Edwards D, Ukaegbu A, Rockcliffe F, Osafo N. Culturally Competent Strategies for Recruitment and Retention of African American Populations into Clinical Trials. Clin Transl Sci. 2015;8(5):460–6.

19. Moreno G, Mangione CM, Meza CE, Kwon I, Seeman T, Trejo L, et al. Perceptions from latino and african american older adults about biological markers in research. Ethn Dis. 2015;25(3):355–62.

20. Williams MM, Scharff DP, Mathews KJ, Hoffsuemmer JS, Jackson P, Morris JC, et al. Barriers and facilitators of African American participation in Alzheimer disease biomarker research. Alzheimer Dis Assoc Disord. 2010;24 Suppl:S24–9.

21. Bonner GJ, Darkwa OK, Gorelick PB. Autopsy recruitment program for African Americans. Alzheimer Dis Assoc Disord. 2000;14(4):202–8.

22. Barnes LL. Biomarkers for Alzheimer Dementia in Diverse Racial and Ethnic Minorities-A Public Health Priority. JAMA Neurol. 2019;76(3):251–3.

23. C. N. Sampling knowledge: the hermeneutics of snowball sampling in qualitative research. Int J Soc Res Methodol. 2009;11(4):327–44.

24. Ajzen I. The theory of planned behavior. Organizational Behavior and Human Decision Processes. 1991;50(2):179–211.

25. Scrimshaw SCM HE. Rapid assessment procedures for nutrition and primary health care: Anthropological approaches to improving programme effectiveness. Los Angeles: : UCLA Latin American Center. ; 1987.

26. Holdsworth LM, Safaeinili N, Winget M, Lorenz KA, Lough M, Asch S, et al. Adapting rapid assessment procedures for implementation research using a team-based approach to analysis: a case example of patient quality and safety interventions in the ICU. Implement Sci. 2020;15(1):12.

27. Lincoln Y, E. G. Naturalistic Inquiry;. Beverly Hills, CA: Sage Publications Inc.; 1985. pp. 334–41. p. 28.

28. Braun V, Clarke V. Using thematic analysis in psychology. Qualitative Research in Psychology,. 2006;3:77–101.

29. Largent EA, Harkins K, van Dyck CH, Hachey S, Sankar P, Karlawish J. Cognitively unimpaired adults’ reactions to disclosure of amyloid PET scan results. PLoS One. 2020;15(2):e0229137.

30. Largent EA, Bhardwaj T, Abera M, Stites SD, Harkins K, Lerner AJ, et al. Disclosing Genetic Risk of Alzheimer’s Disease to Cognitively Unimpaired Older Adults: Findings from the Study of Knowledge and Reactions to APOE Testing (SOKRATES II). J Alzheimers Dis. 2021;84(3):1015–28.

31. Roberts JS, Ferber R, Blacker D, Rumbaugh M, Grill JD, Advisory Group on Risk Evidence Education for D. Disclosure of individual research results at federally funded Alzheimer’s Disease Research Centers. Alzheimers Dement (N Y). 2021;7(1):e12213.

32. Langlois CM, Bradbury A, Wood EM, Roberts JS, Kim SYH, Riviere ME, et al. Alzheimer’s Prevention Initiative Generation Program: Development of an APOE genetic counseling and disclosure process in the context of clinical trials. Alzheimers Dement (N Y). 2019;5:705–16.

33. McCarthy S, Chinman M, Mitchell-Miland C, Schutt RK, Zickmund S, Ellison ML. Peer specialists: Exploring the influence of program structure on their emerging role. Psychol Serv. 2019;16(3):445–55.

34. Chinman M, McCarthy S, Mitchell-Miland C, Bachrach RL, Schutt RK, Ellison M. Predicting Engagement With Mental Health Peer Specialist Services. Psychiatr Serv. 2019;70(4):333–6.

35. Ali-Faisal SF, Colella TJ, Medina-Jaudes N, Benz Scott L. The effectiveness of patient navigation to improve healthcare utilization outcomes: A meta-analysis of randomized controlled trials. Patient Educ Couns. 2017;100(3):436–48.

36. Freeman HP, Rodriguez RL. History and principles of patient navigation. Cancer. 2011;117(15 Suppl):3539–42.

37. Esparza A, Calhoun E. Measuring the impact and potential of patient navigation. Cancer. 2011;117(S15):3535–6.

38. Eliacin J, Fortney SK, Rattray NA, Kean J. Patients’ and caregivers’ perspectives on healthcare navigation in Central Indiana, USA after brain injury. Health Soc Care Community. 2021.

39. Natale-Pereira A, Enard KR, Nevarez L, Jones LA. The role of patient navigators in eliminating health disparities. Cancer. 2011;117(S15):3541–50.

