## Supplementary material for "Factors influencing Blacks and Whites’ participation in Alzheimer’s disease biomarker research": Table 1

| **Table 1. Participants’ Demographic** | Total (N=26)  n, % |
| --- | --- |
| Gender  Male  Female | 11 (42.3%)  15 (57.7%) |
| Age*  55-59  60-69  70-79  80+ | 4 (16.7%)  10 (41.7%)  8 (33.3%)  2 (8.3%) |
| Race  White  Black  Multi | 10 (38.5%)  15 (57.7%)  1 (3.8%) |
| Hispanic  Non-Hispanic* | 0 (0%)  23 (100%) |
| Education*  HS/GED^1^  Some college/2 years  4 years college degree  >4 years college | 2 (9.1%)  5 (22.7%)  7 (31.8%)  8 (36.4.0) |
| Veteran  Non-Veteran | 6 (23.1%)  20 (76.9%) |
| * Has missing values.  ^1^GED: General Education Development | |
